# Impact of accurate initial discharge planning and in-patient transfers of care on discharge delays: a retrospective cohort study

**DOI:** 10.1101/2024.10.11.24315330

**Authors:** Dan Burns, Chris Duckworth, Carlos Lamas Fernandez, Rachael Leyland, Mark Wright, Matthew Stammers, Michael George, Michael Boniface

## Abstract

**Objective:** To investigate the association between initial discharge planning and transfers of in-patient care with discharge delay. To identify operational changes which could expedite discharge within the Discharge to Assess (D2A) model.

**Design:** Retrospective cohort study.

**Setting:** University Hospital Southampton NHS Foundation Trust (UHSFT).

**Participants:** All adults (≥18 years) who registered a hospital spell in UHSFT between 1 January 2021 and 31 December 2022 (n = 258,051 spells). Individuals were followed from hospital admission through to discharge. Data includes demographics, comorbidities, operational information (ward changes, handovers) and discharge information (estimated discharge date, D2A pathway).

**Main outcome measures:** The primary outcome was discharge delay, defined as the number of days between the final estimated discharge date and the actual discharge date. Odds ratio analysis was used to assess the impact of initial discharge planning accuracy (D2A pathway), number of ward moves, and number of in-speciality handovers on the outcome, adjusting for demographic and patient complexity factors.

**Results:** Out of 65,491 spells, 10,619 had an initial planned pathway that did not match the final discharge pathway, with 7,790 of these spells (75.1%) recording a discharge delay. Conversely, 10,216 of 54,872 spells (18.6%) where the initial pathway matched the final pathway recorded a discharge delay (adjusted odds ratio 2.72 (95% CI 2.55 – 2.91)). Ward moves and in-specialty handovers were also associated with increased likelihood of discharge delay, with adjusted odds ratio 1.25 (1.23 – 1.28) per ward move and 1.17 (1.14 – 1.20) per in-specialty handover.

**Conclusions:** This study finds a strong association between inaccurate initial discharge plans and in-patient transfers of care with discharge delay, after controlling for patient complexity and acuity. This highlights the need to consider how initial plans, and in-patient transfers affect discharge planning. Given the lead-times for organising onward care, operational inefficiencies are most impactful for patients eventually discharged on pathways with higher planning complexity.

**Key messages:** What is already known on this topic

- Several factors are known to influence discharge delay including age, whether the spell was elective, and patient complexity.
- Discharge planning has been historically difficult to characterise.

What this study adds

- A robust measurement of the accuracy of discharge planning.
- Highlights the importance of considering the impact of initial discharge plans on the planning process.

How this study might affect research, practice or policy

- Encourage a higher level of consideration when suggesting an initial Discharge to Assess pathway to reduce errors in planning down the line.
- Ensure that relevant discharge information is communicated when patients are transferred between wards and care teams.

## Introduction

The UK National Health Service (NHS) is under the most severe pressure in its 75-year history. In NHS England alone, 7.6 million patients are waiting for treatment, with up-to 2.5 million patients waiting more than a year for elective care^1^. Bed availability remains at critical levels, with occupancy consistently exceeding 93% through winter 2023/24^2^. Contributing to this pressure is the large number of patients remaining in hospital despite no longer meeting the criteria to reside. As of January 2024, 14,436 patients a day on average remained in hospital despite being ready to leave, with delays to discharge steadily rising over the last 3 years (30% higher relative to December 2021)^3^.

Delays to discharge lead to worse outcomes for patients, including physical and cognitive deconditioning and increased risk of hospital-acquired infection^4-8^. Delays also imperil other patients who either experience long trolley waits in the emergency department or are unable to be admitted for urgent elective care^4,9^. Despite 85% of hospital in-patients being discharged without additional support, the most common reason for discharge delay is the requirement for onward social care (over 65% of all spells^7^). The transfer from hospital to social care can be complex. It can be delayed due to capacity problems in the workforce and onward care services, lack of patient information to plan onward care, and inefficiencies in communication or discharge process. For patients in need of onward care, any delay in assessment, choice, or access prevents discharge from hospital^6^.

Discharge planning, designed to mitigate delays to discharge, is the process of setting up an onward package of care for patients in the hospital. This requires the hospital care team to assess and predict the needs of the patient pre-, during, and post-treatment. With social care capacity issues causing a significant lead time on care organisation, delays to discharge are possible if plans cannot be made in a timely manner. Despite this, patient complexity (e.g., age, comorbidities, number of episodes and ward/specialty transfers) greatly impedes clinical ability to make informed and timely predictions of onward care, particularly when the patient’s condition or care needs change abruptly during a hospital spell. Furthermore, planned discharge pathway changes throughout the patient’s stay result in a need to restart the planning process, potentially leading to delays in discharge.

Despite the large-scale impact of discharge delay on the NHS^10,11^, few studies have been conducted to understand its drivers in a hospital setting and are often targeted at specific patient subgroups such as vascular surgery^12^, hip fractures^13^, paediatric intensive care^14^, and trauma^15^ patients. One study has been carried out for admission from an emergency department finding that demographic and arrival mode are determinants of a delayed transfer of care^16^. Meta-analyses on the impact of discharge planning on length of stay have been carried out and reveal small reductions in length of stay overall^17,18^. Despite this, there is a notable lack of standardisation in the definition of discharge planning^19^. A robust definition of planning quality is therefore missing.

In this study, we define a way to measure initial planning quality through the Discharge to Assess model of care^20,21^. Using this, we investigate how discharge delay varies by initial discharge planning, care organisation through the spell and inpatient complexity. We look to quantify how the accuracy of initial planning (i.e., the ability to predict onward care needs) impacts the likelihood and magnitude of discharge delay. As secondary measures, we also consider the impact of two operational circumstances: transfer of care between clinical teams within specialty and ward changes. Throughout, we control for patient complexity by quantifying factors which significantly reduce clinical ability to create accurate onward care plans, and how these complex care needs may be harder to organise.

## Methods

### Data sources and study design

This cohort study is based on routinely collected data at University Hospitals Southampton Foundation Trust (UHSFT). Information, including patient demographics and hospital spells, was extracted directly from the UHSFT’s systems. Hospital spell data details the admission date, discharge date, consultant episodes within the spell, the consultant’s specialty, and ICD10 codes associated with the spell^22^. Each hospital spell is made up of one or more consultant episodes, which is defined as a continuous period where one or more consultants within a particular speciality have leadership over the care of the patient. Each episode contains information about the length of the episode, the associated care speciality, and ICD10 codes recording the reason for the stay and any comorbidities or complications that occurred during the episode. Each spell has a dominant episode (and therefore dominant specialty of care), which is defined as the most resource-intensive episode by the NHS Healthcare Resource Grouper^23^. The database also contains information about which wards the patient stayed in during their spell.

Discharge planning information is extracted from the UHSFT’s Application Express (APEX) system. APEX is an audit log of changes to each patient’s discharge plan with associated timestamps. It contains information about their discharge pathway as assessed by the discharge team, estimated discharge date (EDD), and identified onward care needs. The pathways are characterised by the corresponding numeric pathway in the UK’s Discharge to Assess model of care^20,21^. In this care model, organisation of onward care, ranging from short-term rehabilitation and reablement to more permanent long-term arrangements, are organised into four discharge pathways:

- **Pathway 0** reflects a simple discharge case or a restart of existing care packages.
- **Pathway 1** involves discharge to the usual place or residence with additional support, such as reablement, therapy, or longer-term at-home packages of care.
- **Pathway 2** involves temporary bed-based settings such as in-patient rehabilitation, mental health, or hospice beds with short term service or respite placements.
- **Pathway 3** indicates a permanent care home or long-term bed-based care setting, typically a new admission to a long-term care facility or home, or a return to a pre-existing care home placement.

The pathway reflects a scale of planning complexity: from pathway 0 with minimal planning, to pathway 3 having the highest planning complexity for patients with multiple onward care needs. As the discharge planning process evolves, the pathway and EDD fields are updated and timestamped accordingly. Therefore, there can be multiple pathways and EDD reassessments in each spell record. The final recorded Discharge to Assess pathway represents the actual route through which the patient was discharged, with the final EDD representing the date that a patient is due to be medically fit for discharge.

### Study population

We include all hospital spells concerning adult patients (>= 18 years old) between 1 January 2021 and 31 December 2022. The inclusion criteria were hospital spells that 1) had discharge planning information recorded, 2) the patient is alive at the point of discharge, and 3) had complete demographics and ICD10 codes. We exclude hospital spells which had a rare dominant specialty or admission type, i.e., when there were fewer than 100 spells with that dominant specialty or admission type.

### Outcome measurement

The primary outcome of interest is whether a discharge delay occurred within a given hospital spell. At UHSFT, the local policy indicates that the EDD should be frozen when a patient is medically fit for discharge. This allows for an accurate representation of discharge delay through a comparison of the expected discharge by care teams and the actual discharge date. Therefore, we define discharge delay as the difference between the actual discharge date and the last reported EDD (in days) within the hospital spell. If the EDD is after the discharge date, we set the discharge delay to be 0 days. The EDD is updated on the day the patient is discharged for a subset of spells. In this instance, we take the second-to-last reported EDD (if available) as the estimate. We binarise the discharge delay outcome for odds ratio analysis, with 0 (False) representing a discharge delay of 0 days and 1 (True) for a discharge delay > 0 days.

### Covariates

To describe patient complexity, we extract the number of comorbidities based upon those used in the Charlson Comorbidity Index (CCI) from each hospital spell. We also extract the speciality of the dominant episode (henceforth referred to as the dominant speciality), the number of unique specialities involved in a spell, and whether the spell was elective. The age of the patient at hospital admission is also extracted. We also extract whether the spell involved a stay in an Intensive Care Unit (ICU) from the ward stays data.

To describe operational and planning complexity, we extract the number of ward moves a patient undertakes during a spell and the number of times a spell contained an in-specialty handover between consultant teams. An in-speciality handover is defined as a transfer of care responsibility between consultants of the same speciality (e.g. a consultant in general medicine transferring the patient’s care to another consultant in general medicine).

To describe initial planning, we extract the initial and final recorded Discharge to Assess pathway and create a binary variable indicating whether these pathway records match. In cases where there was only one pathway entry for the patient, the initial pathway and final pathway match by default.

### Statistical analyses

We calculated descriptive counts of each covariate category for the final selected cohort, stratified by whether the spell had a recorded discharge delay. To understand potential cohort selection effects, we compute descriptive statistics regarding the length of stay for the spells that were excluded due to a lack of planning information to explore differences between these cohorts. We performed odds ratio analysis using logistic regression to estimate the effect of operational and planning complexity variates on the risk of discharge delay. We built three models per operational variate (i.e., initial pathway does not match final, number of ward moves, number of in-specialty handovers): 1) a crude model without adjustments; 2) a model with only operational variates (the three covariates of interest, and final pathway); 3) and a fully adjusted model taking into account remaining covariates describing patient complexity. The results are presented as odds ratios (ORs) with corresponding 95% confidence intervals. We check the model quality by assessing the Area Under the Receiver Operating Characteristic curve (AUROC) to ensure that the model has adequate explanatory power with confidence intervals calculated using bootstrapping. To ensure that changes in operational circumstances during the COVID-19 pandemic are considered, we stratify hospital spells by the year of their admission date, resulting in two sets: those starting in 2021 and 2022, respectively. Analyses were performed using Python 3.9.5.

### Subgroup and sensitivity analyses

We evaluate effect modification by building logistic regression models including an interaction term between the three operational variables of interest and each adjustment variable in the fully adjusted model, resulting in a model per adjustment variable. We calculate the p-value for each interaction term. In the case that interactions were significant, i.e p < 0.001, stratified analyses by age, sex, number of comorbidities, and final pathway were planned. In the stratified analyses, we report the ORs of each group.

## Results

### Descriptive statistics

In Figure 1, we show a diagram describing cohort selection, leaving N=65,491 for studying discharge delay at UHSFT during the study period. In studying the cohort selection effects of removing spells without planning information, we found that 84% of excluded spells had a length of stay less than 1 day, and a further 8% with length of stay between 1 and 2 days. Comparing to the chosen cohort, the 84^th^ percentile of length of stay was 17 days, with the 92^nd^ percentile equal to 23 days. Table 1 provides descriptive statistics of the study cohort, stratified by whether a discharge delay was recorded. The cohort is further stratified by each covariate with raw counts and percentages provided.

**Table 1:**
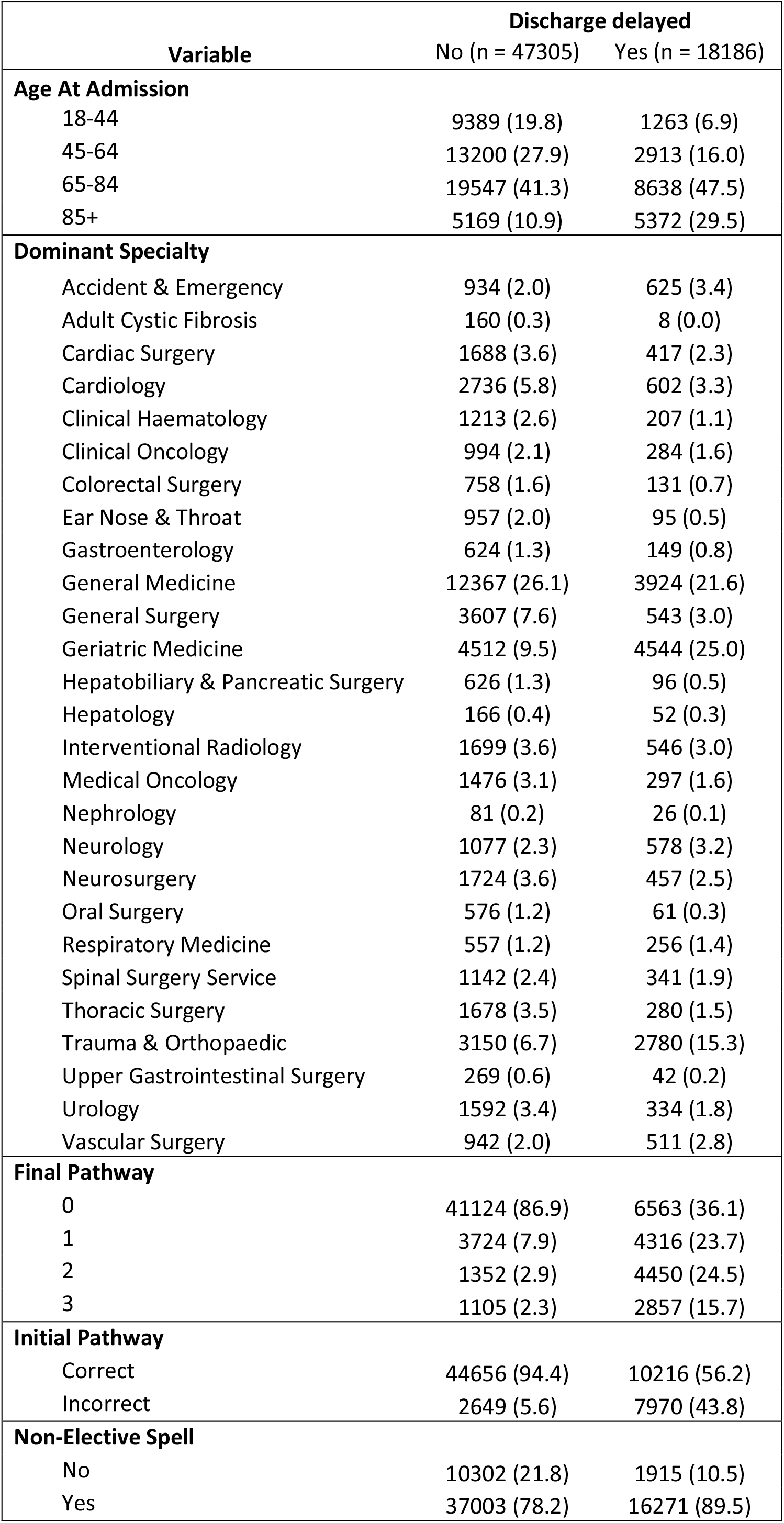
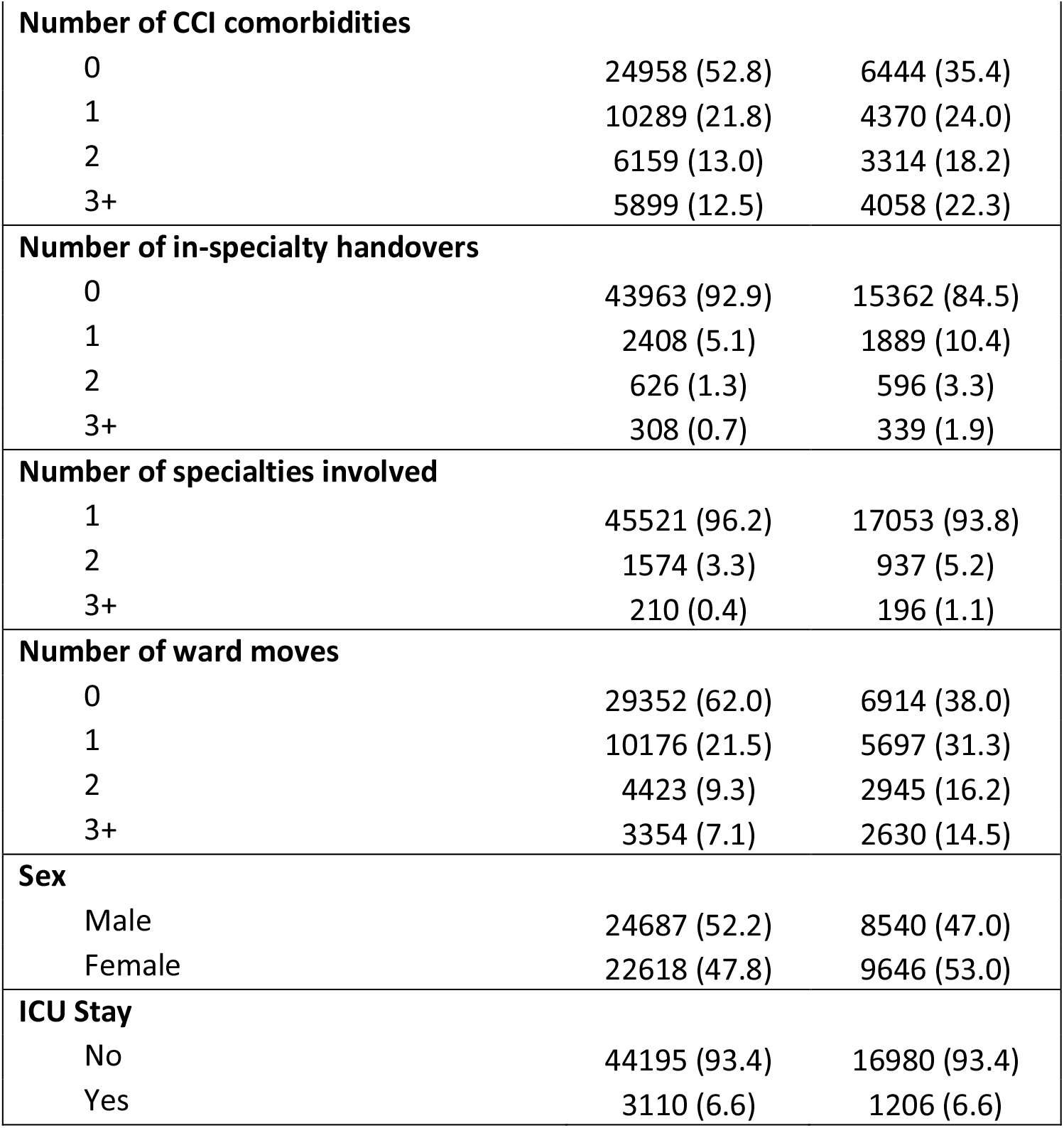
Descriptive statistics of the cohort spells, stratified by whether a discharge delay occurred or not. Values are spell count (percentage).

| Variable | Discharge delayed |  |
| --- | --- | --- |
|  | No (n = 47305) | Yes (n = 18186) |
| **Age At Admission** |  |  |
| 18-44 | 9389 (19.8) | 1263 (6.9) |
| 45-64 | 13200 (27.9) | 2913 (16.0) |
| 65-84 | 19547 (41.3) | 8638 (47.5) |
| 85+ | 5169 (10.9) | 5372 (29.5) |
| **Dominant Specialty** |  |  |
| Accident & Emergency | 934 (2.0) | 625 (3.4) |
| Adult Cystic Fibrosis | 160 (0.3) | 8 (0.0) |
| Cardiac Surgery | 1688 (3.6) | 417 (2.3) |
| Cardiology | 2736 (5.8) | 602 (3.3) |
| Clinical Haematology | 1213 (2.6) | 207 (1.1) |
| Clinical Oncology | 994 (2.1) | 284 (1.6) |
| Colorectal Surgery | 758 (1.6) | 131 (0.7) |
| Ear Nose & Throat | 957 (2.0) | 95 (0.5) |
| Gastroenterology | 624 (1.3) | 149 (0.8) |
| General Medicine | 12367 (26.1) | 3924 (21.6) |
| General Surgery | 3607 (7.6) | 543 (3.0) |
| Geriatric Medicine | 4512 (9.5) | 4544 (25.0) |
| Hepatobiliary & Pancreatic Surgery | 626 (1.3) | 96 (0.5) |
| Hepatology | 166 (0.4) | 52 (0.3) |
| Interventional Radiology | 1699 (3.6) | 546 (3.0) |
| Medical Oncology | 1476 (3.1) | 297 (1.6) |
| Nephrology | 81 (0.2) | 26 (0.1) |
| Neurology | 1077 (2.3) | 578 (3.2) |
| Neurosurgery | 1724 (3.6) | 457 (2.5) |
| Oral Surgery | 576 (1.2) | 61 (0.3) |
| Respiratory Medicine | 557 (1.2) | 256 (1.4) |
| Spinal Surgery Service | 1142 (2.4) | 341 (1.9) |
| Thoracic Surgery | 1678 (3.5) | 280 (1.5) |
| Trauma & Orthopaedic | 3150 (6.7) | 2780 (15.3) |
| Upper Gastrointestinal Surgery | 269 (0.6) | 42 (0.2) |
| Urology | 1592 (3.4) | 334 (1.8) |
| Vascular Surgery | 942 (2.0) | 511 (2.8) |
| **Final Pathway** |  |  |
| 0 | 41124 (86.9) | 6563 (36.1) |
| 1 | 3724 (7.9) | 4316 (23.7) |
| 2 | 1352 (2.9) | 4450 (24.5) |
| 3 | 1105 (2.3) | 2857 (15.7) |
| **Initial Pathway** |  |  |
| Correct | 44656 (94.4) | 10216 (56.2) |
| Incorrect | 2649 (5.6) | 7970 (43.8) |
| **Non-Elective Spell** |  |  |
| No | 10302 (21.8) | 1915 (10.5) |
| Yes | 37003 (78.2) | 16271 (89.5) |
| **Number of CCI comorbidities** |  |  |
| 0 | 24958 (52.8) | 6444 (35.4) |
| 1 | 10289 (21.8) | 4370 (24.0) |
| 2 | 6159 (13.0) | 3314 (18.2) |
| 3+ | 5899 (12.5) | 4058 (22.3) |
| **Number of in-specialty handovers** |  |  |
| 0 | 43963 (92.9) | 15362 (84.5) |
| 1 | 2408 (5.1) | 1889 (10.4) |
| 2 | 626 (1.3) | 596 (3.3) |
| 3+ | 308 (0.7) | 339 (1.9) |
| **Number of specialties involved** |  |  |
| 1 | 45521 (96.2) | 17053 (93.8) |
| 2 | 1574 (3.3) | 937 (5.2) |
| 3+ | 210 (0.4) | 196 (1.1) |
| **Number of ward moves** |  |  |
| 0 | 29352 (62.0) | 6914 (38.0) |
| 1 | 10176 (21.5) | 5697 (31.3) |
| 2 | 4423 (9.3) | 2945 (16.2) |
| 3+ | 3354 (7.1) | 2630 (14.5) |
| **Sex** |  |  |
| Male | 24687 (52.2) | 8540 (47.0) |
| Female | 22618 (47.8) | 9646 (53.0) |
| **ICU Stay** |  |  |
| No | 44195 (93.4) | 16980 (93.4) |
| Yes | 3110 (6.6) | 1206 (6.6) |

**Figure 1:**
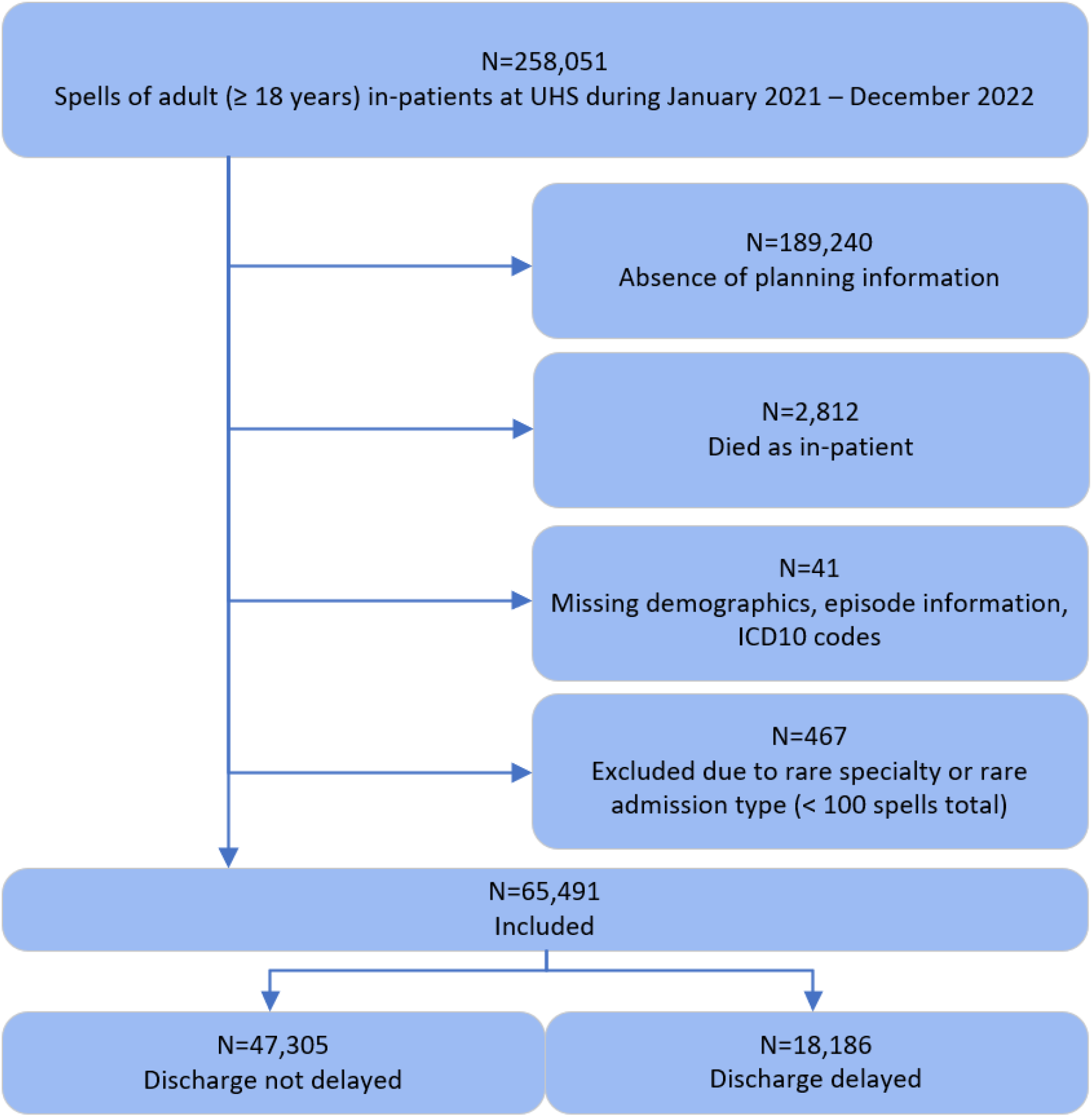
Cohort selection diagram during study period.

In the top panel (A) of Figure 2, we compute the average discharge delay stratified by different covariate values. We find strong correlations (of increasing delay) with non-elective spells, age, number of comorbidities, ward changes, in-speciality handovers, and incorrect initial pathways. Vascular Surgery, Geriatric Medicine and Trauma & Orthopaedic have the highest average delays when stratifying by care speciality, notably all specialities with high likelihood of complex onward care. In the bottom panel (B) of Figure 2, we show the weekly average number of in-patients (dark blue), further stratifying on whether the spell had a discharge delay (light green) or not (teal). We note a general trend of both increased capacity and delayed spells across the study period (01-01-2021 to 31-12-2022).

**Figure 2:**
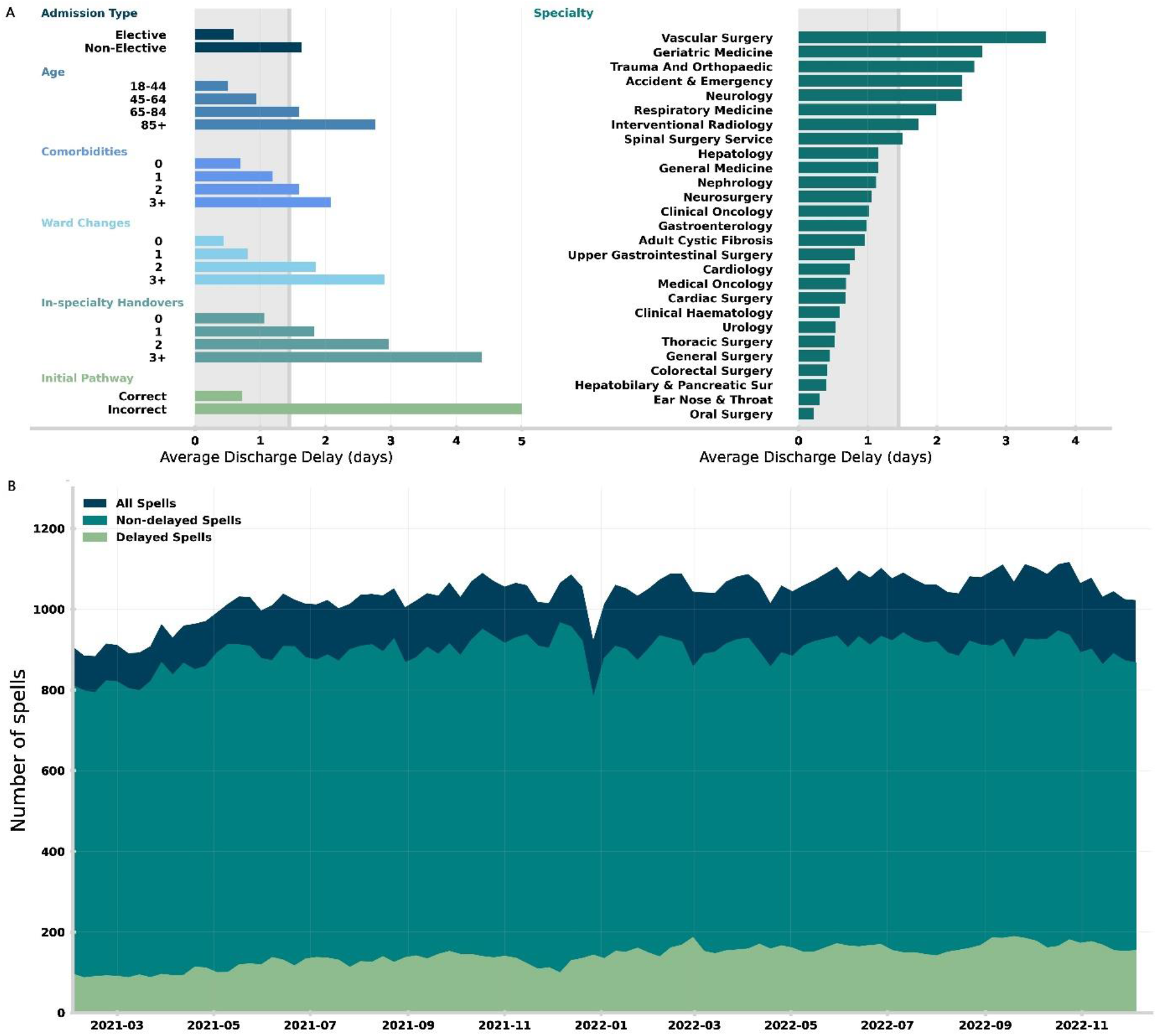
(A) Average discharge delay for hospital spells between 01-01-2021 and 31-12-2022. The grey shading in the background gives the average discharge delay across all spells. Average discharge delay is then computed for spells grouped in different ways: (left, top to bottom) admission type, age at admission, number of registered comorbidities, number of ward changes during the spell, number of in-speciality handovers (i.e., consultant handovers), and whether the initial pathway estimate was correct. (Right) Average discharge delay by dominant speciality, only including specialities with at least 100 registered spells. (B) weekly average of the number of in-patients between 01-01-2021 and 31-12-2022, showing number of in-patients who are delayed with patients who are not delayed.

### Likelihood of discharge delay

Table 2 shows the odds ratios (OR) for our crude, operationally adjusted, and fully adjusted logistic regression models. The fully adjusted logistic regression model achieves an AUROC of 0.823 (95% confidence interval 0.818 – 0.827), indicating that the model has sufficient explanatory power. Inaccurate initial discharge planning (i.e., an initially planned pathway not matching the final pathway) was associated with an increase in the likelihood of discharge delay with adjusted OR 2.72 (2.55 – 2.91). This effect was also observed after stratifying by time, with an adjusted OR of 1.92 (1.73 – 2.14) for spells in 2021 and 2.46 (2.23 – 2.72) for spells in 2022. Each of the secondary operational measures; number of ward moves and in-specialty handovers, were associated with increases in likelihood of discharge delay with adjusted OR 1.25 (1.23 - 1.28) per ward move and 1.17 (1.14 - 1.20) per in-specialty handover. The table of odds ratios for the fully adjusted model is provided in Table S1 in the supplementary materials.

**Table 2:** Odds ratios between initial pathway different to final, number of ward moves, and number of in-specialty handovers, with discharge delay for the crude, operationally adjusted, and fully adjusted models. Odds ratios are quoted as the value of the odds ratio with the 95% confidence interval in brackets.

| Variable | Total spells | Odds ratio |  |  | P value |
| --- | --- | --- | --- | --- | --- |
|  |  | Crude | Operationally adjusted model | Fully adjusted model |  |
| Initial pathway correct | 65,491 | Reference | Reference | Reference |  |
| Initial pathway incorrect |  | 13.15 (12.53 – 13.80) | 2.87 (2.68 – 3.06) | 2.72 (2.55 – 2.91) | <0.001 |
| Stratified by time |  |  |  |  |  |
| 2021 | 32,968 | 13.18 (12.34 – 14.07) | 2.03 (1.83 – 2.26) | 1.92 (1.73- 2.14) | <0.001 |
| 2022 | 32,523 | 13.82 (12.82 – 14.88) | 2.58 (2.34 – 2.85) | 2.46 (2.23 – 2.72) | <0.001 |
| Secondary measures |  |  |  |  |  |
| Number of ward moves | 65,491 | 1.37 (1.35 - 1.39) | 1.26 (1.24 - 1.28) | 1.25 (1.23 - 1.28) | <0.001 |
| Number of in-specialty handovers |  | 1.53 (1.50 - 1.56) | 1.12 (1.09 - 1.15) | 1.17 (1.14 - 1.20) | <0.001 |

### Subgroup and sensitivity analyses

For the initial pathway not matching the final variable, interactions between age, whether the spell was elective, whether the spell included a stay in ICU, number of specialities involved, and final pathway all had p < 0.001. Stratification by D2A pathway revealed that patients discharged on Pathway Patients discharged on Pathway 3 were more affected by an erroneous initial pathway, (OR 4.45 (3.77 - 5.25)), whereas those on Pathway 2 were less affected (OR 1.82 (1.54 - 2.14)),with Pathway 0 (OR 2.72 (2.33 - 3.17)) and Pathway 1 (OR 2.57 (2.30 - 2.87)) remaining similar to the overall OR. For stratification by age, patients were split into four groups for stratification analysis: 18-44, 45-64, 65-84, and 85+. Patients in the 85+ age group had the highest impact from an inaccurate initial pathway (OR 3.23 (2.88 - 3.63)), with the 45-64 age group having the lowest impact (OR 1.82 (1.50 - 2.20)). Stratifications by the number of comorbidities and sex were not carried out as the interaction terms were insignificant.

For ward moves and in-specialty handovers, interactions with age, final pathway, number of specialties involved and whether the spell included a stay in ICU had p < 0.001. Patients discharged on Pathway 3 are most likely to be affected by ward moves (OR 1.41 (1.30 - 1.52)) and in-specialty handovers (OR 1.37 (1.23 - 1.52)). Pathway 1 patients were similar for both ward moves (OR 1.39 (1.32 - 1.45)) and in-specialty handovers (OR 1.39 (1.30 - 1.48)). Patients in the 85+ age group were most affected by ward moves (OR 1.49 (1.143 - 1.56)) and in-specialty handovers (OR 1.20 (1.12 - 1.27)). The odds ratios for the fully adjusted models stratified by D2A pathway and age are provided in Table S2 and S3 respectively in the supplementary materials.

## Discussion

In this study, we found a strong association between the accuracy of an initial pathway assessment and discharge delay, with the odds of discharge delay increasing almost three-fold if the initial discharge pathway assessment is incorrect. We find that patients ultimately discharged on Pathway 3 are most affected by inaccurate initial planning with the odds of discharge delay increasing almost five-fold if the initial pathway is incorrect. We also find that operational processes such as ward changes and in-speciality handovers (i.e., a transfer of care between consultants of the same speciality) have a substantial impact on the likelihood of delay, with three ward changes or four in-speciality transitions doubling the odds of delay. The effects from ward changes and in-speciality handovers remain after correcting for in-patient complexity (e.g., comorbidities, number of care specialities involved in treatment) and is strong for the most acute onward care needs (i.e., Pathway 3). As a result, those with the potentially highest acuity onward care needs should be prioritised when considering operational improvements (e.g., reducing transfer of care).

Since Pathway 3 is a permanent care placement, both local authorities and integrated care boards must be confident and assured that all other pathway options have been exhausted and that the patient’s needs are clear (including a robust mental capacity assessment and wishes are captured). As a result, Pathway 3 requires the most extended lead-times to organise onward care. This has been identified in different patient sub-groups including surgical patients^24^ and trauma patients^15^. Early and accurate identification of potential discharge needs provides discharge teams additional time to organise care and avoid delay. Despite this, early identification of onward discharge needs is fundamentally hard and is exacerbated by extended periods of high bed occupancy.

A significant bottleneck on patient flow through the NHS is the capacity to discharge patients with new onward care needs as they become medically fit. Delays in discharge can derive from inefficiencies to in-hospital processes (e.g., inaccurate prediction of onward care need resulting or delays awaiting test results); however, they are exacerbated by the ability of over-capacity onward care services to accept additional patients. Community services face consistent issues with both staffing and funding, causing significant lead-times for hospital care teams to be able to organise care, especially for care home placements^10^. Despite this, internal processes within hospitals contribute to delay, which should be targeted to help improve patient flow. Efficient and accurate planning in the hospital enables organisation to start earlier and absorb the potential long lead-times on care placement.

There are several possible mechanisms for why transfers of in-patient care contribute to discharge delay. When the transfer occurs, information surrounding the patient’s care must be transferred to the new ward or consultant from their previous care. Information transfer between care teams may often be imperfect, and lack of care continuity potentially contributes to certain information around discharge planning being lost. Specifically in the case of ward changes, system inertia may contribute where patients are transferred to other wards to free up space in acute wards for emergency department patients and, consequently, moved to a lower priority relative to other more unwell patients, leading to less focus on these patients around the initial expected discharge time. Furthermore, periods of increased pressure with high levels of bed occupancy reduce clinical time to accurately assess discharge requirements early and can cause displacement of patients under the same specialty across the hospital^25^. Coupled with unexpected changes to patient conditions or complications from treatment, predicting complex care needs is intrinsically difficult. Clinical decision support tools (e.g., machine learning algorithms) have the potential to help clinical teams better identify patients with complex onward care requirements early. In future work, we demonstrate the ability of an explainable machine learning model to help identify appropriate discharge pathways.

### Strengths and limitations of this study

This study has several strengths. Firstly, the data collection for discharge planning is large and robust due to its usage by complex discharge teams at UHSFT, which itself is the largest hospital (defined by number of beds) in NHS England. The data is digitised and used for national reporting, feeding directly into the statistical snapshot for acute discharge delays aggregated by NHS England. Consequently, the estimated discharge date and discharge pathway updates are accurately recorded. The hospital spell data is also comprehensive as this data is routinely collected and used for national reporting. This study also benefits from direct collaboration between hospital care teams, discharge teams and researchers to provide different perspectives.

This study also has limitations. A key limitation is the absence of planning information for the majority of hospital spells at UHSFT which are not considered in this study. Ingestion of discharge pathway information into APEX only occurs for more complex cases and, consequently, our patient cohort are more complex than average. Despite this, the vast majority of hospital spells that were removed due to lack of discharge planning information were patients with a length of stay less than 1 day. Therefore, many of these are day cases where discharge planning is not required and thus are likely non-complex patients. Other limitations include that this study is single site (i.e., has no other site validating results) and discharge planning in other sites may differ.

## Conclusions

We find that inaccurate initial discharge plans are a significant factor in increasing the likelihood of discharge delay. Clinical decision support tools to correctly assign patients to a discharge pathway may help with accurate and timely initial assessment and may be more important than predicting length of stay. Patients at high risk of requiring complex onward care needs (i.e., short- and long-term intensive care) should be a primary target for improving patient flow due to in-hospital delays in assessment and arrangement of onward care. Patients with complex needs should, wherever possible, remain on a single ward under the same team since these significantly impact the odds ratio of a delayed discharge.

## Ethical Approvals

This study was approved by the University of Southampton’s Ethics and Research Governance Committee (ERGO, and approval was obtained from the Health Research Authority. All methods were carried out following relevant guidelines and regulations associated with the Health Research Authority, and NHS Digital. The research was limited to the use of previously collected, non-identifiable information. Informed consent was waived by the University of Southampton’s Ethics and Research governance committee, University of Southampton, University Road, Southampton, SO17 1BJ, United Kingdom and the Health Research Authority, 2 Redman Place, Stratford, London, E20 1JQ, UK. Data was pseudonymised (and, where appropriate, linked) before being passed to the research team. The research team did not have access to the pseudonymisation key.

## Data Availability

Due to data protection constraints the dataset used in this study is not publicly available. However, it will be made available upon reasonable request.

## Competing interests

The authors have no completing interests.

## Funding

This report includes independent research supported partly by the National Institute for Health Research Applied Research Collaboration Wessex and Health Data Research UK, an initiative funded by UK Research and Innovation, Department of Health and Social Care (England) and the devolved administrations, and leading medical research charities. The views expressed in this publication are those of the author(s) and not necessarily those of the National Institute for Health Research or the Department of Health and Social Care.

## Authors’ contributions

M.B, C.L.F, D.B, C.D conceived the research question. D.B performed the analysis of data and modelling with support from M.B, C.L.F, C.D. D.B wrote the first draft of the manuscript. All other authors contributed to the first and future iterations of the manuscript. C.D, C.L.F, D.B, M.B, M.G, M.S, had access to all data, with C.D. and M.J.B. verifying the quantitative data. M.B obtained ethical and governance approvals. M.G. and M.S. managed the data extraction at UHS. R.L and M.W provided clinical insight.

## Acknowledgements

This report includes independent research funded by the National Institute for Health Research Applied Research Collaboration Wessex. The views expressed in this publication are those of the author(s) and not necessarily those of the National Institute for Health Research or the Department of Health and Social Care. This work was supported by Health Data Research UK, an initiative funded by UK Research and Innovation, Department of Health and Social Care (England) and the devolved administrations, and leading medical research charities. We thank the Southampton Emerging Therapies and Technologies (SETT) Centre at UHS for support with data access and insight.

## Notes

### Competing Interest Statement

The authors have declared no competing interest.

### Summary of Updates

Supplemental files added with references in main text. Minor updates to methods sections, including statistical analysis and subgroup and sensitivity analyses sections to enhance clarity. Addition of age groups used in stratification to subgroup and sensitivity analyses results section, and addition of result for ward moves in the age-stratified analysis.

