## Supplementary Materials for "Impact of accurate initial discharge planning and in-patient transfers of care on discharge delays: a retrospective cohort study"

| Variable | Odds ratio (95% CI) |
|---|---|
| **Intercept** | 0.03 (0.03 - 0.04) |
| **Age** | 1.01 (1.01 - 1.01) |
| **Non-Elective Spell** | 1.21 (1.13 - 1.30) |
| **Sex** | 1.00 (0.96 - 1.04) |
| **Final pathway (Reference: Pathway 0)** | |
| Pathway 1 | 4.38 (4.11 - 4.66) |
| Pathway 2 | 6.12 (5.61 - 6.69) |
| Pathway 3 | 7.15 (6.55 - 7.81) |
| **Dominant Specialty (Reference: General Medicine)** | |
| Accident & Emergency | 1.22 (1.07 - 1.39) |
| Adult Cystic Fibrosis | 0.47 (0.22 - 1.00) |
| Cardiac Surgery | 1.03 (0.89 - 1.20) |
| Cardiology | 0.93 (0.83 - 1.04) |
| Clinical Haematology | 0.99 (0.83 - 1.18) |
| Clinical Oncology | 1.27 (1.08 - 1.49) |
| Colorectal Surgery | 1.03 (0.83 - 1.27) |
| Ear Nose & Throat | 0.80 (0.63 - 1.01) |
| Gastroenterology | 0.59 (0.47 - 0.73) |
| General Surgery | 0.87 (0.77 - 0.97) |
| Geriatric Medicine | 1.14 (1.06 - 1.23) |
| Hepatobilary & Pancreatic Surgery | 0.94 (0.74 - 1.20) |
| Hepatology | 1.15 (0.80 - 1.67) |
| Interventional Radiology | 1.37 (1.20 - 1.55) |
| Medical Oncology | 0.99 (0.85 - 1.15) |
| Nephrology | 0.96 (0.57 - 1.61) |
| Neurology | 1.82 (1.59 - 2.08) |
| Neurosurgery | 1.80 (1.58 - 2.05) |
| Oral Surgery | 0.82 (0.62 - 1.10) |
| Respiratory Medicine | 1.27 (1.06 - 1.53) |
| Spinal Surgery Service | 1.91 (1.65 - 2.22) |
| Thoracic Surgery | 1.29 (1.11 - 1.50) |
| Trauma And Orthopaedic | 2.50 (2.29 - 2.72) |
| Upper Gastrointestinal Surgery | 0.84 (0.58 - 1.21) |
| Urology | 1.11 (0.96 - 1.28) |
| Vascular Surgery | 3.14 (2.75 - 3.58) |
| **Initial pathway (Reference: Correct)** | |
| Incorrect | 2.72 (2.55 - 2.91) |
| **Number of specialties involved** | 1.04 (1.00 - 1.09) |
| **Number of in-speciality handovers** | 1.17 (1.14 - 1.20) |
| **Number of specialties involved** | 1.04 (1.03 - 1.05) |
| **ICU Stay** | 1.09 (0.99 - 1.21) |
| **Number of ward moves** | 1.25 (1.23 - 1.28) |

*Table S1: The odds ratios for the fully adjusted model for each variable with 95% confidence intervals in parentheses.*

| Variable | Stratified Odds Ratios (95% CI) by D2A Pathway | | | |
|---|---|---|---|---|
| | **Pathway 0** | **Pathway 1** | **Pathway 2** | **Pathway 3** |
| **Intercept** | 0.04 (0.03 - 0.05) | 0.04 (0.03 - 0.07) | 0.40 (0.22 - 0.73) | 0.02 (0.01 - 0.05) |
| **Age** | 1.01 (1.01 - 1.01) | 1.02 (1.02 - 1.03) | 1.01 (1.01 - 1.02) | 1.03 (1.02 - 1.03) |
| **Non-Elective Spell** | 1.28 (1.19 - 1.38) | 1.18 (0.89 - 1.57) | 1.03 (0.77 - 1.39) | 1.85 (1.06 - 3.22) |
| **Sex (Reference: Male)** | | | | |
| Female | 1.06 (1.00 - 1.12) | 0.92 (0.84 - 1.02) | 0.99 (0.87 - 1.12) | 0.81 (0.69 - 0.96) |
| **Dominant Specialty (Reference: General Medicine)** | | | | |
| Accident & Emergency | 1.50 (1.23 - 1.83) | 0.61 (0.47 - 0.79) | 0.72 (0.51 - 1.02) | 1.41 (0.90 - 2.21) |
| Adult Cystic Fibrosis | 0.41 (0.17 - 1.00) | -- | -- | -- |
| Cardiac Surgery | 1.23 (1.04 - 1.44) | 0.28 (0.10 - 0.83) | 0.54 (0.24 - 1.22) | 0.10 (0.02 - 0.56) |
| Cardiology | 1.03 (0.90 - 1.18) | 0.79 (0.56 - 1.12) | 1.15 (0.73 - 1.81) | 0.84 (0.45 - 1.57) |
| Clinical Haematology | 1.09 (0.89 - 1.33) | 0.72 (0.41 - 1.27) | 0.60 (0.34 - 1.04) | 2.02 (0.80 - 5.11) |
| Clinical Oncology | 1.30 (1.07 - 1.59) | 0.80 (0.48 - 1.32) | 1.47 (0.91 - 2.39) | 1.91 (0.83 - 4.41) |
| Colorectal Surgery | 1.23 (0.99 - 1.55) | 0.22 (0.06 - 0.83) | 0.77 (0.30 - 1.94) | 0.49 (0.09 - 2.55) |
| Ear Nose & Throat | 0.88 (0.68 - 1.13) | 1.00 (0.43 - 2.32) | 0.49 (0.19 - 1.22) | 0.29 (0.08 - 0.99) |
| Gastroenterology | 0.58 (0.44 - 0.78) | 0.59 (0.34 - 1.02) | 0.66 (0.32 - 1.39) | 0.63 (0.28 - 1.41) |
| General Surgery | 1.01 (0.89 - 1.14) | 0.62 (0.45 - 0.85) | 0.42 (0.28 - 0.63) | 0.66 (0.36 - 1.20) |
| Geriatric Medicine | 1.35 (1.19 - 1.53) | 0.71 (0.61 - 0.84) | 0.94 (0.74 - 1.18) | 0.93 (0.71 - 1.22) |
| Hepatobilary & Pancreatic Surgery | 1.11 (0.86 - 1.43) | 0.10 (0.02 - 0.47) | -- | 1.28 (0.20 - 8.36) |
| Hepatology | 0.99 (0.63 - 1.58) | 3.39 (1.04 - 11.01) | -- | -- |
| Interventional Radiology | 1.80 (1.56 - 2.08) | 0.71 (0.47 - 1.08) | 0.56 (0.37 - 0.86) | 0.70 (0.36 - 1.34) |
| Medical Oncology | 0.93 (0.77 - 1.12) | 1.53 (0.94 - 2.50) | 0.71 (0.46 - 1.09) | 2.01 (0.97 - 4.15) |
| Nephrology | 0.65 (0.28 - 1.51) | 1.71 (0.56 - 5.23) | 1.17 (0.31 - 4.43) | 0.33 (0.02 - 6.96) |
| Neurology | 2.04 (1.72 - 2.41) | 1.71 (1.21 - 2.40) | 1.04 (0.72 - 1.49) | 1.85 (1.11 - 3.07) |
| Neurosurgery | 2.06 (1.79 - 2.38) | 0.70 (0.35 - 1.43) | 0.76 (0.45 - 1.27) | 3.26 (1.63 - 6.50) |
| Oral Surgery | 0.81 (0.58 - 1.12) | 1.44 (0.59 - 3.53) | 1.07 (0.20 - 5.77) | 0.58 (0.07 - 4.76) |
| Respiratory Medicine | 1.24 (0.97 - 1.60) | 1.62 (1.06 - 2.47) | 1.06 (0.61 - 1.86) | 1.31 (0.64 - 2.68) |
| Spinal Surgery Service | 2.11 (1.79 - 2.48) | 1.85 (1.08 - 3.19) | 0.81 (0.49 - 1.32) | 2.70 (1.01 - 7.23) |
| Thoracic Surgery | 1.47 (1.25 - 1.72) | 1.28 (0.63 - 2.60) | 0.54 (0.27 - 1.07) | 2.32 (0.60 - 9.04) |
| Trauma And Orthopaedic | 3.14 (2.82 - 3.49) | 2.14 (1.72 - 2.65) | 1.07 (0.85 - 1.36) | 2.77 (1.96 - 3.92) |
| Upper Gastrointestinal Surgery | 0.89 (0.59 - 1.35) | 1.64 (0.54 - 4.99) | 0.26 (0.05 - 1.24) | -- |
| Urology | 1.23 (1.04 - 1.46) | 0.85 (0.59 - 1.24) | 1.21 (0.67 - 2.20) | 1.04 (0.53 - 2.05) |
| Vascular Surgery | 3.86 (3.35 - 4.45) | 1.04 (0.62 - 1.76) | 1.30 (0.73 - 2.31) | 0.85 (0.27 - 2.63) |
| **Initial pathway (Reference: Correct)** | | | | |
| Incorrect | 2.72 (2.33 - 3.17) | 2.57 (2.30 - 2.87) | 1.82 (1.54 - 2.14) | 4.45 (3.77 - 5.25) |
| **Number of specialties involved** | 0.98 (0.92 - 1.03) | 1.35 (1.20 - 1.51) | 1.00 (0.87 - 1.14) | 1.42 (1.18 - 1.71) |
| **Number of in-speciality handovers** | 1.08 (1.04 - 1.12) | 1.39 (1.30 - 1.48) | 1.18 (1.08 - 1.29) | 1.37 (1.23 - 1.52) |
| **Number of comorbidities** | 1.06 (1.04 - 1.08) | 1.03 (1.00 - 1.06) | 0.97 (0.93 - 1.01) | 1.05 (1.00 - 1.11) |
| **ICU Stay** | 1.24 (1.11 - 1.39) | 1.07 (0.69 - 1.65) | 0.68 (0.49 - 0.94) | 0.79 (0.48 - 1.29) |
| **Number of ward moves** | 1.22 (1.20 - 1.25) | 1.39 (1.32 - 1.45) | 1.29 (1.22 - 1.36) | 1.41 (1.30 - 1.52) |

*Table S2: The odds ratios for the fully adjusted model, with patients stratified by final discharge to assess pathway (95% confidence intervals in parentheses). Cases where the odds ratio confidence interval has a width larger than 10 have been suppressed.*

| Variable | Stratified Odds Ratios (95% CI) by Age Group | | | |
|---|---|---|---|---|
| | **18-44** | **45-64** | **65-84** | **85+** |
| **Intercept** | 0.05 (0.04 - 0.07) | 0.05 (0.04 - 0.06) | 0.06 (0.05 - 0.07) | 0.05 (0.03 - 0.08) |
| **Non-Elective Spell** | 1.44 (1.17 - 1.77) | 1.24 (1.08 - 1.41) | 1.19 (1.07 - 1.31) | 1.08 (0.84 - 1.39) |
| **Sex (Reference: Male)** | | | | |
| Female | 0.84 (0.74 - 0.96) | 0.96 (0.87 - 1.05) | 1.09 (1.03 - 1.16) | 0.97 (0.88 - 1.07) |
| **Final pathway (Reference: Pathway 0)** | | | | |
| Pathway 1 | 1.96 (1.41 - 2.72) | 5.25 (4.48 - 6.16) | 5.36 (4.89 - 5.86) | 3.91 (3.46 - 4.43) |
| Pathway 2 | 3.80 (2.30 - 6.25) | 8.08 (6.22 - 10.50) | 6.54 (5.77 - 7.40) | 5.72 (4.86 - 6.73) |
| Pathway 3 | 3.66 (2.61 - 5.14) | 7.83 (6.14 - 9.99) | 9.08 (7.92 - 10.42) | 6.92 (5.90 - 8.13) |
| **Dominant Specialty (Reference: General Medicine)** | | | | |
| Accident & Emergency | 2.01 (1.38 - 2.92) | 1.33 (0.97 - 1.83) | 1.07 (0.88 - 1.31) | 1.66 (1.19 - 2.32) |
| Adult Cystic Fibrosis | 0.35 (0.13 - 0.98) | 0.78 (0.22 - 2.83) | 0.44 (0.03 - 6.54) | -- |
| Cardiac Surgery | 1.28 (0.76 - 2.18) | 0.84 (0.62 - 1.12) | 1.24 (1.01 - 1.52) | 0.96 (0.38 - 2.43) |
| Cardiology | 1.36 (0.90 - 2.04) | 0.92 (0.72 - 1.19) | 0.88 (0.76 - 1.03) | 1.56 (1.10 - 2.22) |
| Clinical Haematology | 1.11 (0.66 - 1.86) | 1.15 (0.84 - 1.57) | 0.83 (0.64 - 1.08) | 2.56 (1.35 - 4.85) |
| Clinical Oncology | 1.82 (1.01 - 3.26) | 1.37 (1.02 - 1.85) | 1.19 (0.95 - 1.48) | 2.15 (1.07 - 4.34) |
| Colorectal Surgery | 1.40 (0.79 - 2.48) | 1.39 (0.97 - 2.00) | 0.97 (0.70 - 1.33) | 0.50 (0.20 - 1.21) |
| Ear Nose & Throat | 0.97 (0.66 - 1.42) | 0.78 (0.47 - 1.28) | 0.65 (0.43 - 0.99) | 1.35 (0.63 - 2.85) |
| Gastroenterology | 0.94 (0.60 - 1.47) | 0.52 (0.34 - 0.79) | 0.54 (0.38 - 0.76) | 0.91 (0.44 - 1.89) |
| General Surgery | 0.99 (0.78 - 1.26) | 0.99 (0.80 - 1.23) | 0.84 (0.70 - 1.00) | 1.03 (0.70 - 1.51) |
| Geriatric Medicine | -- | 1.74 (0.57 - 5.33) | 1.13 (1.01 - 1.26) | 1.77 (1.34 - 2.34) |
| Hepatobiliary & Pancreatic Surgery | 1.39 (0.71 - 2.70) | 1.20 (0.78 - 1.85) | 0.88 (0.63 - 1.23) | 0.47 (0.12 - 1.84) |
| Hepatology | 1.44 (0.68 - 3.07) | 1.20 (0.68 - 2.11) | 1.05 (0.55 - 2.01) | -- |
| Interventional Radiology | 1.81 (1.28 - 2.57) | 1.48 (1.17 - 1.87) | 1.22 (1.02 - 1.47) | 2.61 (1.59 - 4.28) |
| Medical Oncology | 1.18 (0.74 - 1.87) | 1.41 (1.09 - 1.82) | 0.78 (0.63 - 0.97) | 1.52 (0.62 - 3.72) |
| Nephrology | -- | 1.29 (0.58 - 2.87) | 0.78 (0.37 - 1.64) | -- |
| Neurology | 2.13 (1.42 - 3.21) | 1.77 (1.33 - 2.36) | 1.83 (1.52 - 2.22) | 2.96 (1.97 - 4.44) |
| Neurosurgery | 1.98 (1.46 - 2.68) | 1.78 (1.41 - 2.23) | 1.84 (1.49 - 2.26) | 3.10 (1.70 - 5.66) |
| Oral Surgery | 0.84 (0.49 - 1.43) | 1.09 (0.65 - 1.84) | 0.58 (0.34 - 0.99) | 1.87 (0.77 - 4.56) |
| Respiratory Medicine | 1.69 (0.99 - 2.88) | 1.62 (1.16 - 2.25) | 1.04 (0.81 - 1.34) | 1.92 (0.67 - 5.48) |
| Spinal Surgery Service | 1.45 (1.03 - 2.04) | 2.27 (1.76 - 2.93) | 2.00 (1.57 - 2.54) | 3.35 (1.84 - 6.10) |
| Thoracic Surgery | 1.02 (0.62 - 1.68) | 1.47 (1.09 - 1.97) | 1.21 (0.99 - 1.48) | 4.26 (2.34 - 7.75) |
| Trauma And Orthopaedic | 2.83 (2.29 - 3.51) | 2.82 (2.35 - 3.38) | 2.47 (2.18 - 2.81) | 3.35 (2.46 - 4.57) |
| Upper Gastrointestinal Surgery | 0.60 (0.14 - 2.60) | 1.17 (0.67 - 2.05) | 0.69 (0.40 - 1.19) | -- |
| Urology | 0.89 (0.58 - 1.39) | 1.13 (0.82 - 1.55) | 1.13 (0.92 - 1.38) | 2.21 (1.43 - 3.41) |
| Vascular Surgery | 3.14 (1.74 - 5.66) | 4.66 (3.73 - 5.82) | 2.68 (2.23 - 3.22) | 3.15 (1.76 - 5.62) |
| **Initial pathway (Reference: Correct)** | | | | |
| Incorrect | 2.06 (1.50 - 2.84) | 1.82 (1.50 - 2.20) | 2.68 (2.43 - 2.96) | 3.23 (2.88 - 3.63) |
| **Number of specialties involved** | 0.98 (0.87 - 1.11) | 1.01 (0.92 - 1.10) | 1.11 (1.04 - 1.18) | 1.01 (0.87 - 1.17) |
| **Number of in-speciality handovers** | 1.16 (1.07 - 1.26) | 1.15 (1.09 - 1.22) | 1.17 (1.12 - 1.22) | 1.20 (1.12 - 1.27) |
| **Number of comorbidities** | 1.06 (0.99 - 1.12) | 1.08 (1.04 - 1.11) | 1.05 (1.03 - 1.07) | 1.00 (0.97 - 1.03) |
| **ICU Stay** | 1.66 (1.31 - 2.10) | 1.20 (1.01 - 1.42) | 1.02 (0.88 - 1.19) | 0.65 (0.38 - 1.11) |
| **Number of ward moves** | 1.17 (1.11 - 1.24) | 1.23 (1.19 - 1.27) | 1.22 (1.19 - 1.26) | 1.49 (1.43 - 1.56) |

*Table S3: The odds ratios for the fully adjusted model, with patients stratified by age group (95% confidence intervals in parentheses). Cases where the odds ratio confidence interval has a width larger than 10 have been suppressed.*
